# Voluntary commitment of medical students during the COVID-19-pandemic

**DOI:** 10.1101/2020.11.21.20236067

**Authors:** Lorenz Leopold Mihatsch, Mira von der Linde, Franziska Knolle, Benjamin Luchting, Konstantinos Dimitriadis, Jens Heyn

## Abstract

**Introduction:** Due to the spreading of COVID-19, one key challenge was to reduce potential staff shortages in the healthcare sector. Besides a reactivation of retired healthcare workers, medical students were considered to support this task. We herein analysed the commitment of medical students during the COVID-19-pandemic as well as their burdens and anxieties.

**Methods:** Via an online survey, medical students in Germany were asked regarding these themes. The survey period took place during a two-week period in April and Mai 2020. 1241 questionnairs were included in the analysis.

**Results:** 67.9% [65.3; 70.5] of the participants reported that they volunteered during the pandemic. Simultaneously, 88.9% [86.9; 90.5] of the participants stated to be against a compulsory recruitment in this context. Students who volunteered showed a significantly lower anxiety index than students, who did not volunteer. Concerns about transferring the virus to patients or relatives were significantly higher than concerns about the students’ own infection.

**Conclusion:** The results of this survey study suggest that the majority of students are working voluntary during the pandemic. They declared mostly that they had adequate opportunities to protect themselves. Finally, their biggest fear was to infect other patients, relatives, or friends.

## Introduction

At the beginning of the COVID-19 pandemic, the knowledge about the virus was very limited. Early data from China suggested that health care workers have an increased risk of infection with the virus [1]. Due to the spreading of COVID-19 and growing numbers of infected persons, the question arose how to cover shortages of staff within the health care sector [2]. During the search for potential practitioners, two specific groups were identified namely retired healthcare workers and medical students to support this task [3]. However, the reactivation of retired staff was problematic, as it became clear that the risk of severe infection with COVID-19 increases with age [4].

Therefore, medical students moved into the main focus as workers of a heavily polluted healthcare sector [3]. This idea is not fundamentally new. There are two prominent examples from the past century when medical students were involved in fighting and overcoming a pandemic/epidemic [5]. During the Spanish flu (1918), medical students at the University of Pennsylvania took care of patients [6]. Approximately four decades later (1952), medical students were tasked with the manual ventilation of tracheostomised patients during the Danish polio epidemic [7]. Based on the experiences of the past, it is not surprising that several countries tried to accelerate the graduation in the field of medicine and to prepare freshly graduated physicians to treat patients with COVID-19 [5]. Neither in the past nor during the actual pandemic have medical students been analysed relating to their commitment in a pandemic or their anxieties of a potential infection with the virus for instance. We therefore addressed these questions in our questionnaire: (i) Are the students willing to work voluntary? (ii) What are their biggest burdens and anxieties? (iii) Do they have enough possibilities to protect themselves and have they already had a covid infection?

## Methods

### Ethics

The questioning was performed in accordance with the Declaration of Helsinki and it was approved on 19.05.2020 by the institutional Review Board of the Ludwig-Maximilians University of Munich (20-458 KB). All participants were enlightened with respect to purpose, objectives and handling of the data which were captured in the questionnaire. Informed consents were given by all participants.

### Survey design

During the period of two weeks (25.05. - 07.06.2020), we systematically questioned medical students in Germany via an online platform (provided by Technical University of Munich) [8]. The questions dealt with different aspects of the student’s commitment during the COVID-19 pandemic. We systematically contacted all student representatives of medical faculties in Germany in order to share the link to the online platform with the students. Furthermore, we also systematically contacted the medical students via social media. We calculated our sample size to be at least n = 383 (with 95% - confidence interval, 5% error and population size of 100.000 medical students in Germany).

### Participants

In total, 1407 medical students participated in the questioning, 1241 were included in the final analysis. Reasons for excluding participants from the analysis were indicating to study in Bremen or Brandenburg in two cases (there are no medical faculties), stating diverse as gender (there is no statistical inference possible with a single person), and 163 students during their final year (commitment is obligatory in that final year). There is no significant difference in the commitment of students from northern (n_north_ = 485 participants) and southern (n_south_ = 746 participants) Germany (p>0.99). We therefore assume our sample to be representative of the medical students in Germany.

### Index of anxiety

Adapted to the anxiety definition from Asendorf and Caspar we developed three items for the purpose of measuring trait anxiety.^9^ These items were measured on a ten-grades Likert scale. As a requirement of offsetting the answers to an index value, we premised an internal test consistency of Cronbach’s Alpha > 0.8 and a selectivity > 0.3 (Item-Scale-Correlation) between the respective questions. Those requirements were fulfilled for the questions about anxiety (alpha: 0.81, selectivity: min. 0.64). The index was calculated regarding to

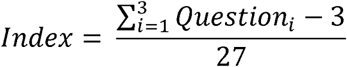

Therefore, the index score takes values between of the interval [0, 1]. 1 is the maximum and 0 the minimal index score. For measuring empathy by four items we maximally scored a Cronbach’s Alpha = 0.62, which did not meet our premise.

### Statistical analysis

Statistical analysis was performed using R Version 3.6.0 (R core team (2019), Vienna, Austria). For group differences we used the Pearsons’s Chi^2^ test for categorical variables and the Wilcox rank sum test for quasi-steady variables. 95% confidence intervals are given in square brackets.

## Results

The proportion of female participants in this survey was 75.8%, with male participants 24.2% respectively. Female medical students had a median age of 24 (with IQR from 22 to 27). Male students were slightly younger with a median age of 23 (with IQR from 24 to 26). 67.9% [65.3; 70.5] of the students declared that they supported the health care system voluntary during the COVID-19 pandemic. Out of these, the proportion of male students was higher compared to female students (75.0% [69.7; 79.8] versus 65.7% [62.5; 68.7], p=0.003; figure 1A). 88.9% [86.9; 90.5] of the interviewed students argued against an obligation of medical students to assist during the COVID-19 pandemic (figure 2).

**Figure 1:**
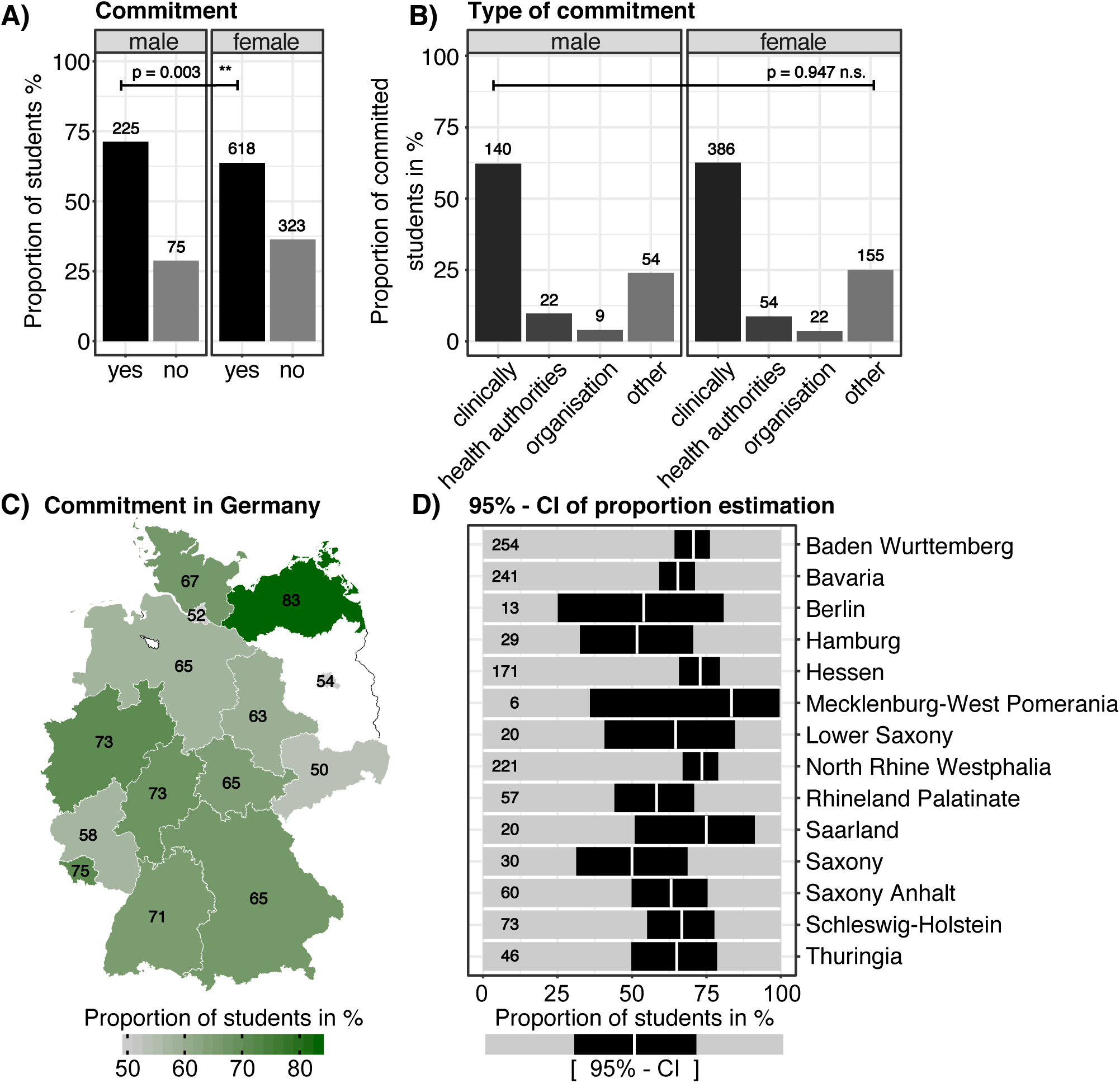
Commitment of medical students during the COVID-19 pandemic. (A) Relative proportion of medical students in Germany with a voluntary commitment. Numbers above columns indicate the absolute number of students in each group. Test for statistical significance was performed using the chi-squared-test. (B) Relative proportion of different types of commitment. The type “organization” refers to organizational work in the medical faculty. Test for statistically significant differences between male and female students was performed using the chi-squared-test. (C) Estimation of the proportion of committed students in German federal states. (D) In the states Bremen and Brandenburg, there are no medical faculties. The number in each state refers to the proportion in % of voluntarily committed students in each state. (E) Visualization of the 95% confidence intervals of the estimated proportions of committed students in German states. White vertical bars depict the estimated proportions in %, black boxes refer to the confidence interval, numbers at the beginning of each bar indicate the number of students questioned in the corresponding state.

**Figure 2:**
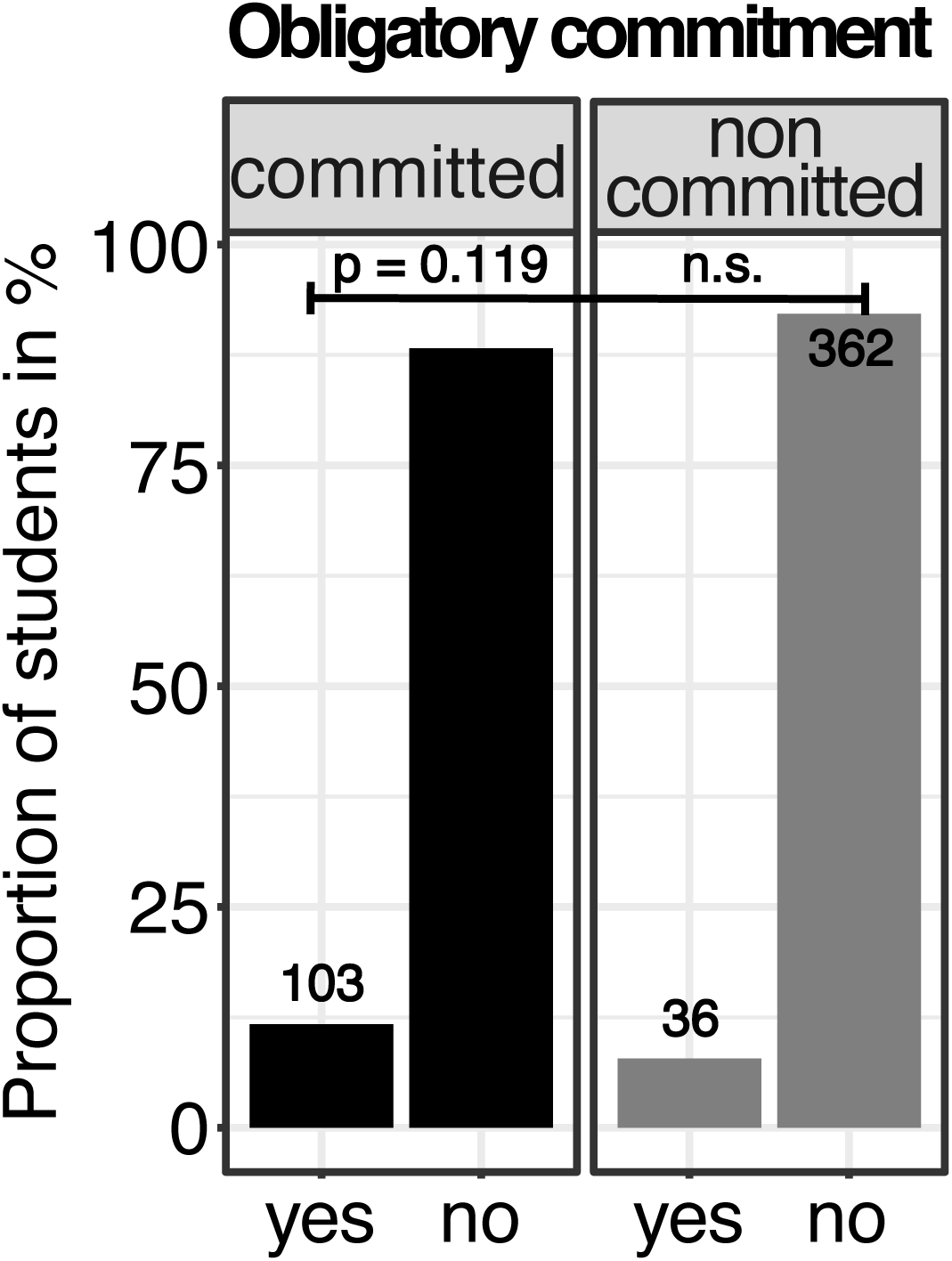
Attitude towards an obligation. (A) Opinion of German medical students concerning an obligatory commitment during the pandemic in relative numbers. Numbers above columns indicate the absolute number of students in each group. Test for statistical significance between committed and non-committed students was performed using the chi-squared-test.

Concerning the type of commitment, there was no significant difference between the sexes. Both, male and female students engaged most dominantly in a clinical context (figure 1B). Slight regional differences between states in Germany were not significant and do not suggest any geographic correlation. The proportions of voluntarily committed students ranged from 50 - 83.3% (figure 1C – D).

In order to address possible differences between committed and non-committed students we hypothesized that intrinsic traits, like anxiety and empathy, may influence the willingness to engage oneself. Indeed, the calculated anxiety index showed a respective relationship. Non-committed medical students showed significantly higher index values, than committed students (p<0.001; figure 3A). Overall female students showed a significantly higher anxiety index, then male students (p<0.001).

**Figure 3:**
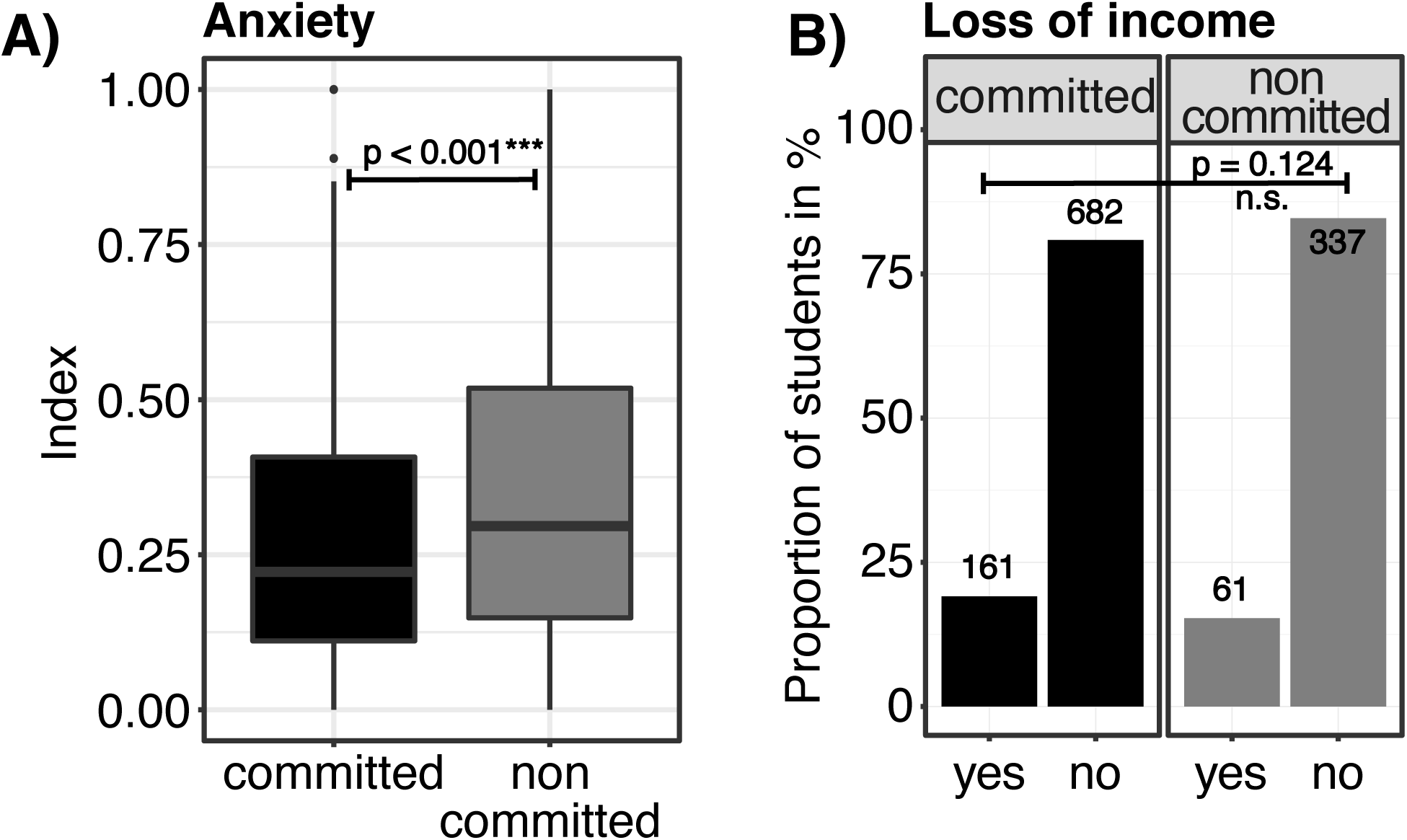
Factors influencing the commitment. (A) Comparison of the anxiety index of committed versus non-committed students. Calculation of the index is given in the method section. Test for significant differences was determined using the Wilcoxon rank-sum test. (B) Relative proportion of medical students, that faced a loss of income during the COVID-19 pandemic. Test for statistical significance between committed and non-committed students was performed using the chi-squared-test.

The feeling of being able to protect oneself against COVID-19, measured on a Likert scale from 1 (low) to 10 (high), had a median value of 7 for male and 6 for female students (p<0.001). It was lowest in female students, that had no voluntary commitment (median: 5, p<0.001). Besides intrinsic traits we also hypothesized financial difficulties as extrinsic motivational factors influencing one’s commitment. Such financial difficulties occurred in 17.9% [15.8; 20.1] of all participants but were not significantly different between the two groups (p=0.13; figure 3B).

As opposed to anxiety as a personal trait, we directly addressed concerns with COVID-19 infections. It turned out that such concerns were significantly lowest to one’s own person. It was higher in terms of infecting other patients and highest for infecting people in one’s personal environment (figure 4A). Based on self-disclosure, the overall infection rate for the students was estimated to be 1.5% [0.9; 2.4].

**Figure 4:**
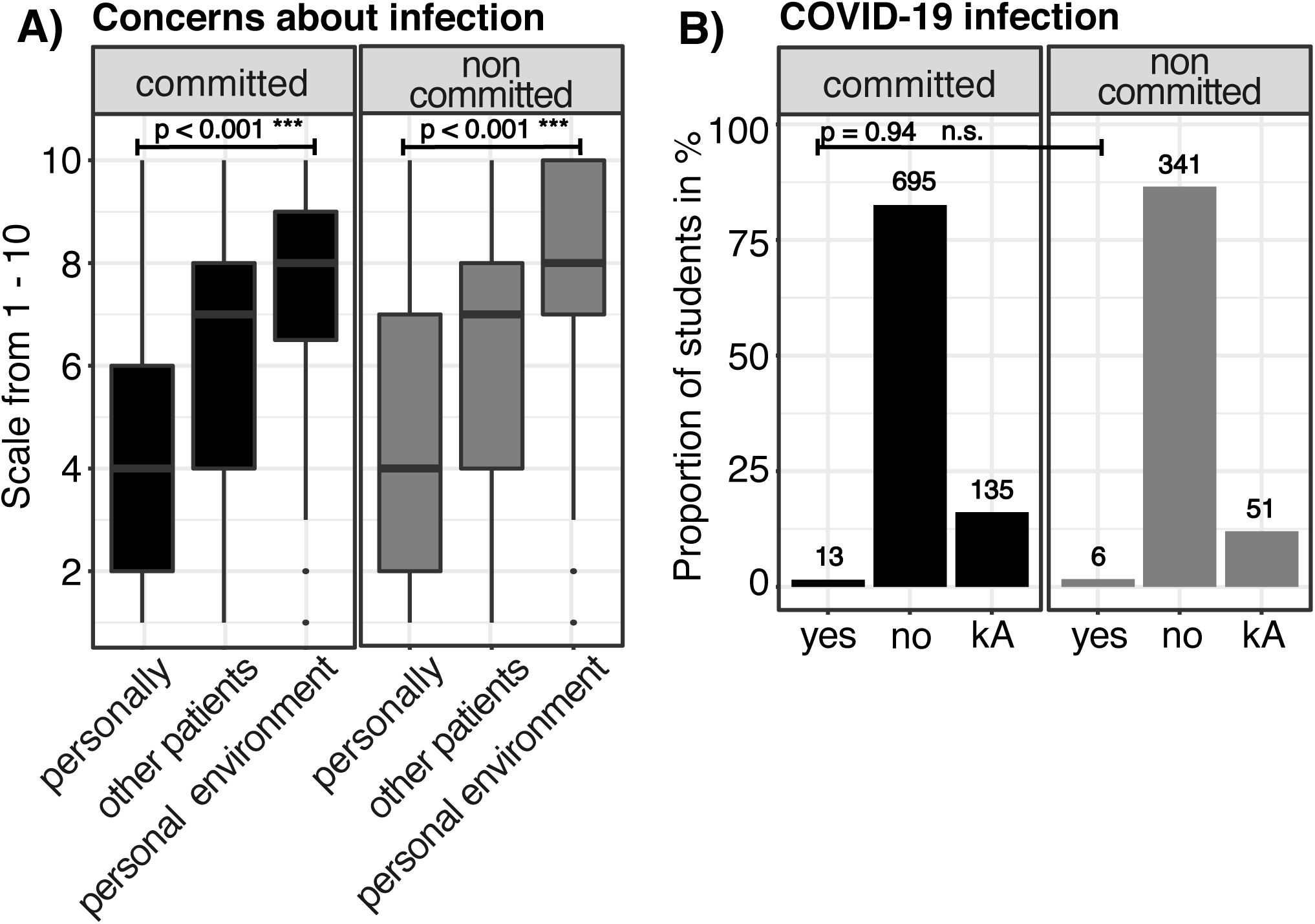
Risk of infection. (A) Fear of committed and non-committed medical students of being infected themselves, of unwillingly infecting patients and people of their personal environment. Measured on a Likert scale from 1 (low fear) to 10 (high fear). (B) Relative proportion in % of students, which have been infected with COVID-19 based on self-disclosure. Test for statistical significance between committed and non-committed students was performed using the chi-squared-test.

## Discussion

With our study, we are able to provide first insights how a pandemic and a possible commitment in fighting the pandemic impacts on the lives, thoughts and feelings of medical students. Furthermore, we could show that the majority of medical students committed voluntarily and refused an obliged commitment. Interestingly, students were less concerned about their own infection with COVID-19, than about the infection of patients and relatives. Overall less than 2% of all participants declared that they have or had been infected with COVID-19.

During the past, medical students showed often that they favor the concept of volunteering. They are willing to campaign for that. This was last proven, when medical students protested against a reform of the licensing regulations, which intended an obligatory rotation at a general practitioner during the last year of medical school [10]. Therefore, it is not surprising that the vast majority of students refuses an obligatory commitment in times of a pandemic. Since the majority of students stated to support the health system voluntary during the pandemic, an obligatory commitment seems to be not necessary.

In order to prevent further spread of the virus worldwide several businesses, factories, stores and restaurants have been closed. Consequently, an enormous number of jobs were lost or lead to short-term work, which resulted in a loss of income [11,12]. Our data reflects this process also for medical students. About every fifth student was affected by such loss of income and put them under financial stress [11,12]. Therefore, the governmental decision of a financial support for students in Germany was definitely in the spirit of the students [13].

Maner et al. [14] previously showed that young adults have a high level of trait anxiety leads to avoid risky situations. Our results fit well into that scheme. We found an inverse connection: higher trait anxiety was connected to less willingness to volunteer during the pandemic and to be exposed to the risk of being infected. In a survey that analyzed the psychic condition of 1000 Greek students from various disciplines, students showed a significant increase in anxiety and depression during the pandemic. Similarly, an increase in questioning the meaning of life was documented [15]. In another work which was performed in China by Coa et al. [16] the level of anxiety of medical students was analyzed. 75% of the students stated a “normal” level of anxiety while only 3% reported a high level of anxiety. Furthermore, the level of anxiety was depending on various factors, like economic factors, prolonged study duration, changes in everyday life and social structure during the pandemic [16]. The data of our survey confirmed this observed overall low level of anxiety. However, the level of anxiety was significantly higher in female than in male medical students. The level of anxiety was especially high in non-committed students.

Wearing adequate protective gear is necessary for self-protection and effective in preventing further viral spread [17]. At the beginning of the pandemic a bottleneck in the delivery of protective gear was reported repetitively [18]. It was therefore of interest to pick up on this central issue. The majority of medical students reported to be able to sufficiently protect themselves. One could speculate that the opinion on non-sufficient protection increases anxiety in this group and explains this lower commitment.

One possible approach to further increase the commitment could be via improved educational work on safety measures and risks. This education should also include the information that voluntary commitment does not necessarily mean to work directly with COVID-19 patients. It would also be supportive if medical students support the regular health care operations. Alternatively, they could support the healthcare departments or to organize online medical education.^5^ This would be an excellent option for students who are willing to be committed but worry about infection with COVID-19.

Our study has some limitations:

- Data collection during a defined time reflects the impressions at this specific time point. A dramatic increase in infected patients or the availability of a vaccine/therapy may have an impact on these results
- After our systematic consultation of the student representatives of German medical faculties, we had no influence on further distribution of the link. This may lead to minor estimation biases. As we do not see significant differences in the proportion of committed students between northern and southern Germany, we still assume our study population to be representative of medial students in Germany.

## Conclusion

The results of this survey study suggest that the majority of students are working voluntary during the pandemic. They decline an obligatory commitment for students when fighting the pandemic. Most of them declared that they had adequate opportunities to protect themselves. Their infection rates were comparable with the German population. Finally, their biggest fear was to infect other patients, relatives, or friends.

## Data Availability

Data can be shared on request for personal use.

## Author contribution

LLM, ML, FK, and JH designed the study. LLM, BL, and KD collected the data. LLM performed the statistical analysis. All authors were involved in the interpretation of the data. LLM and JH wrote the manuscript. ML, FK, BL, and KD were involved in drafting the manuscript and revising it critically for important intellectual content. All authors read and approved the final manuscript.

